# There’s an App for That: Exploring the Market for Contraceptive Fertility Tracking Apps in the Philippines

**DOI:** 10.1101/2023.07.31.23293317

**Authors:** K Danna, DM Harris, CW Rothschild, B Brogaard, E LaCroix, M Paudel

## Abstract

For generations, women have relied on fertility awareness methods (FAM) to plan and prevent pregnancy; for over a decade, many have been aided by digital tools to do so. New Contraceptive Fertility Tracking Apps (CFTAs)--that are backed by clinical efficacy trials to support their effectiveness as contraception--have the potential to enhance method choice and offer users a unique contraceptive option, but there is little evidence to inform the decisions around expanding access, particularly in low-and middle-income countries (LMICs). We conducted a mixed methods study with quantitative online surveys (n=1600) and qualitative components (n=36) to explore the potential appeal of and demand for a hypothetical CFTA in one such market, the Philippines. Interest in using a CFTA was high among our internet-engaged, urban study population, with 83.9% “definitely” or “probably” interested in using. Across demographic profiles, respondents perceived the appeal of the method as “natural” and “convenient.” A majority were willing to pay for the method, though notably at a price (5.20 USD) below that of currently available CFTAs. We explore various factors to be considered before bringing a method like this to new markets, including equity constraints and the prevalence of other period-tracking apps not intended as contraception.

## Background

Leveraging the potential impact of self-care interventions to improve the health of individuals and strengthen health systems could be transformational; following the global COVID-19 pandemic, the relevance of self-care solutions to improve women’s reproductive health access and autonomy is especially evident.^1^ While people have long taken health decisions into their own hands, today’s technological advances mean that reliable health information and effective self-care solutions are easier than ever to find. When they are accessible, acceptable, and affordable, self-care interventions can enable a health system to meet the needs of more people, overcome health system challenges, and increase choice and autonomy.^2^

In the context of reproductive health, self-care solutions to family planning (FP) are among the most promising approaches to ensure that men and women have the tools they need to achieve their reproductive health goals. Expanding method choice with contraceptive options that meet clients’ diverse health needs can improve contraceptive prevalence.^3^ The addition of self-care methods can contribute toward universal health coverage by reducing reliance on overburdened health systems, increasing their efficiency, and overcoming some of the supply-side barriers common with new method introduction, such as the need for provider training and other supply chain constraints.^4^ Contraceptive options like self-administered injectables, for example, can provide an excellent choice for women seeking privacy, efficacy, and ease of use; but a contraceptive application (app) for your smartphone goes one step further — by removing the visit to the health provider, health facility, or pharmacy altogether.

Period tracking apps are not new; the first versions of the apps in use today came on the market in 2013.^5^ Health apps, particularly those targeting women, are a rapidly growing industry and period tracking apps are among the most popular type of app in this category,^6^ with even major tech companies adding period tracking features to their app offerings. While most period tracking apps on the market are not based on Fertility Awareness Methods (FAMs)^7, 8^ and lack the capacity to provide complete and accurate data for reliable pregnancy prevention (and are not explicitly intended for this purpose), a few have evolved into apps that *can* effectively be used as contraception.^9, 10^ These apps, which we will refer to as Contraceptive Fertility Tracking Apps (CFTAs) in order to distinguish them as a contraceptive method category distinct from other period tracking apps, offer users a modern-day tool to employ FP methods that have been available in analog form for over a century. Generally, CFTAs build on the foundation of other FAMs such as the Billings, Creighton, Two-day, or Standard Days methods. Users track their menstrual cycle days and/or changes to their body (temperature and/or cervical mucus) and input this information into the app. The combination of these user inputs plus more complex data/modeling and proprietary algorithms used by the apps mean that CFTAs can more accurately predict an individual user’s fertile window compared to other FAMs. CFTAs, unlike basic period trackers, also offer behavioral instructions for preventing pregnancy on fertile days. Recently, two CFTAs have undergone clinical efficacy trials and have, for the first time, been granted clearance from the US Food and Drug Administration (FDA) to be promoted as contraception.^11, 12^ This market is likely to continue growing.

Like other short-acting contraceptive methods, CFTAs’ effectiveness relies on users’ behaviors: those who wish to avoid pregnancy are instructed to use a barrier method or abstain from sex on fertile days. With typical use, CFTAs are shown to be 92-93% effective.^13, 14^ CFTAs may appeal to users who desire a natural, hormone-free option that is in their control, easy to use, and does not require visits to a provider or pharmacy to use. From a public health perspective, there is a potential for these apps to provide an accessible self-care option across many settings and to improve the effectiveness of other methods (e.g., when used in combination with condoms, emergency contraception, etc.) to prevent pregnancy. However, there are important limitations — the cost of these apps can be high (over $100/year in the US^15^) and require access to a smartphone. While the emerging evidence around their effectiveness is promising, peer-reviewed research studying CFTAs is relatively sparse compared to other established contraceptive methods.^16–18^ CFTAs are also not reliably effective for users with absent or irregular menstrual cycles or cycle variation.^19, 20^ Lastly, there are some concerns around data privacy, especially considering the intimate nature of the data these apps record and the potential for this information to be sold to advertisers. In more restrictive legal environments regulating reproductive health services, data privacy may be of particular concern.^21^

The Philippines is a promising potential market for CFTAs, as its economy is growing at its fastest rate in decades^22^ and more than half of all adults own or use a smartphone.^23^ The country has seen a steady increase in the use of modern contraceptives, although the modern contraceptive prevalence rate (mCPR) for women of reproductive age remains at a modest 26.8%.^24^ Furthermore, more than 2.5 million married women and 5 million unmarried women in the Philippines want to delay or avoid pregnancy but are not using a modern method of contraception to do so.^25^ Unmet need is highest amongst married adolescents (aged 15-19) and those with no formal education (27.9% and 24.3%, respectively). Additionally, the method mix shows a heavy skew toward oral contraception and female sterilization, which may suggest that a more robust range of method options is needed^26^ to ensure full contraceptive choice.^27^ Furthermore, CFTAs could provide an additional tool to help improve users’ body/health literacy, which could have catalytic impact in a setting where knowledge around pregnancy risk is low. For example, only 1 in 4 women (24%) in the Philippines correctly report that a woman is most at risk of pregnancy if she has intercourse halfway between two menstrual periods, even among women who already use FAM.^28^

A deep understanding of the consumer and market environment is critically important for the introduction of FP products into new markets. The purpose of this study was to contribute to the evidence base on the potential for CFTAs to impact the FP marketplace in low- and middle-income countries (LMIC). This exploratory research project aimed to garner understanding of the potential market and interested population(s) for a hypothetical CFTA. We carried out qualitative and quantitative data collection to explore the following research questions:

- What are the socio-demographic and sexual and reproductive health characteristics of potential CFTA users, including their current contraceptive use behaviors and future intentions to use contraception?
- What characteristics, attributes, and limitations of CFTAs are appealing or unappealing to potential users?
- What is the willingness to pay for CFTAs?

## Methods

This analysis describes the results from two components of a larger parent study, which used a sequential mixed methods design consisting of four parts in total. First, we conducted qualitative friendship pair discussions primarily intended to pretest an informational video introducing a hypothetical CFTA used in the later quantitative data collection phase (described in more detail in Data Collection). Following revisions to the video based on findings from the friendship pair discussions, quantitative online surveys were used to gauge respondents’ reactions to the CFTA presented in the video, including their interest in using the method and willingness to pay for the potential product. A subset of this online survey cohort participated in qualitative in-depth interviews (IDIs) to dive deeper into insights and themes emerging from the quantitative phase. Lastly, a smaller cohort of individuals recruited offline were given a shorter version of the quantitative online survey used to see if any notable differences were evident between the study groups recruited digitally versus offline. While this final survey was not powered to detect significant differences between the groups, insights gained from comparison were used to help to refine assumptions used in subsequent efforts to model the size of the market for CFTAs in the Philippines (model to be published separately). The authors selected this mixed methods approach because it allowed for pretesting and revision of the CFTA informational video and provided the opportunity to seek deeper qualitative insights into quantitative associations or trends. The analyses presented here focus on the quantitative survey and the IDIs, the two research components with results that respond directly to our primary research questions.

### Study Population and Sampling Strategy

The primary audience for a future introduction of a CFTA would be an online/internet-engaged population. As such, we chose the online survey approach to ensure that the survey responses would be representative of this population. Women were eligible for inclusion if they were of reproductive age (18-49), expressed interest in using a contraceptive method to delay or prevent pregnancy now or in the future, and if they own/regularly use a smartphone with internet connectivity. The quantitative survey utilized an online panel recruited and maintained by our local research partner, Nielsen IQ, to identify and recruit potential participants. This online panel has been used regularly to conduct studies across multiple industry sectors and consists of over 1.4 million panelists.

The sample size was calculated to estimate prevalence of the primary outcome, interest in using a CFTA, with a precision of +/-2.5% points. Because data on interest is sparse, sample size calculations estimated interest in CFTA use at 50% to ensure adequate precision.

### Participant Recruitment

An initial roster of panel members for the study was created by filtering for gender, age, and residence (women, 18-49yrs, residing in urban areas). Information on these characteristics was collected and maintained by Nielsen IQ as part of panel membership. From this roster, an e-mail invitation to join the survey was sent randomly to 11,982 women. Participants received a generic invitation used for online surveys that said, “Ladies! This new study is for you! Complete the survey using the link below [Survey Link]”. This invitation did not contain any details about the survey and further details were only provided after accepting the invitation. Panel members who received an email were able to electronically opt-in to complete an electronic survey that included additional eligibility screening and, among those eligible, an online electronic consent process. Those meeting these criteria were allowed to proceed to the online survey questionnaire while those not meeting the eligibility criteria exited the survey process. Of those invited to participate, 3,524 women accepted and were asked consent using the online consent form which included details on the study and what was involved in participation. Among those who accepted the invitation, 30 did not consent to the survey. During the screening process, 1,868 were found to be ineligible based on the study criteria. Of the remaining 1,626 possible participants, 26 started but did not complete the survey questionnaire (Figure 1). This response rate was within the expected range given Nielsen IQ’s prior experience with online surveys.

**Figure 1.**
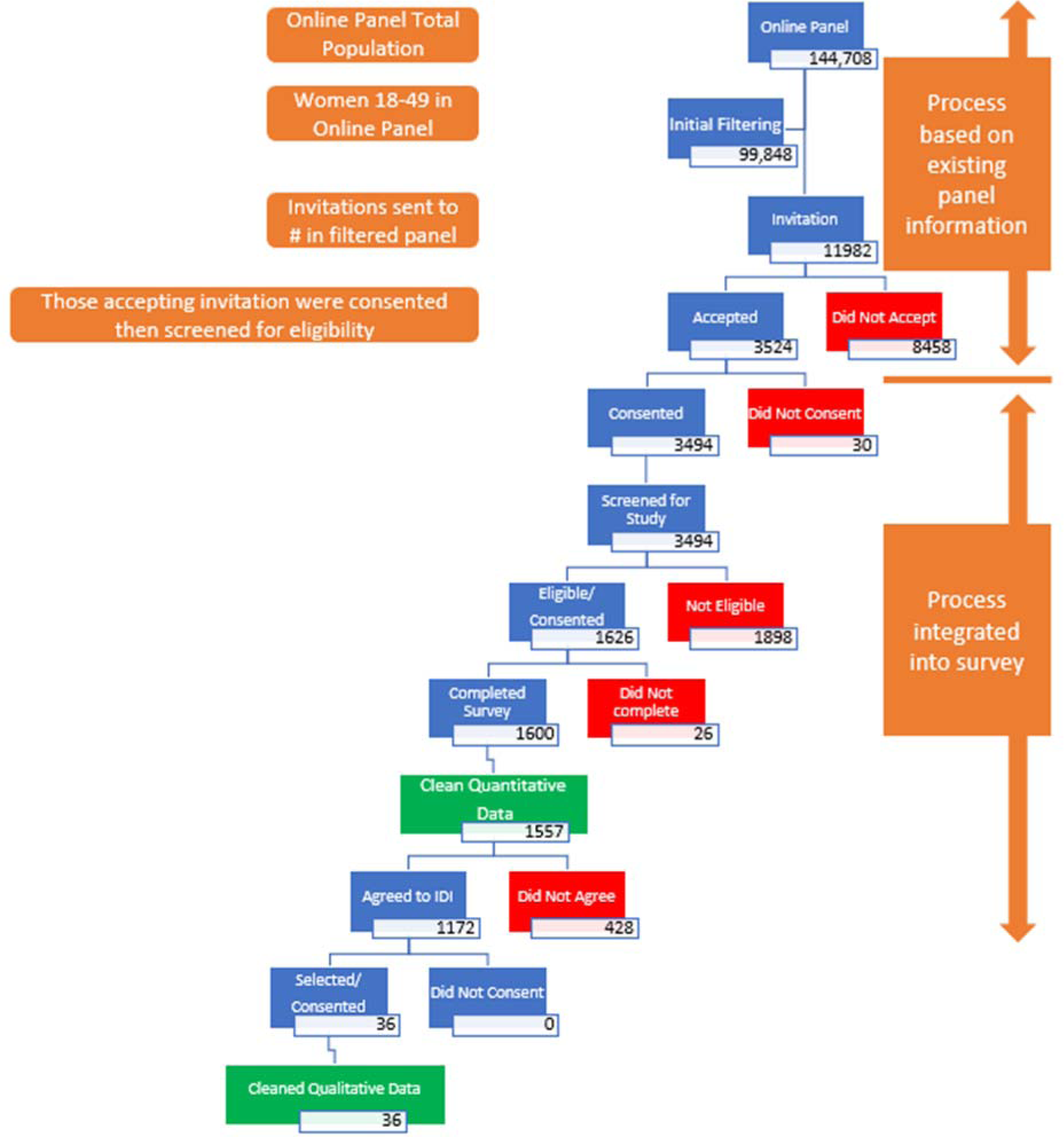
Data Collection Process

At the end of the survey, participants were asked about their interest in participating in a future qualitative IDI. Those who expressed interest constituted the sample frame for the IDI component (1172). Among those, a total of 36 participants, 12 from each age group (18-24, 25-34, and 35-49), were selected randomly for an IDI. All participants consented to the IDI prior to its start.

### Data Collection

The quantitative online survey sought data and insights about participants’ perceptions of a hypothetical CFTA, as presented to them via the informational video (pretested during the friendship pair discussions), and their willingness to use and pay for such an app. The qualitative IDIs sought to gather deeper insights regarding participant preferences and responses, and to understand the reasons behind response trends seen in the quantitative phase.

The quantitative self-administered online survey was conducted using a structured questionnaire. The consent forms and study tools were translated into Tagalog and contained English translations to ensure better understanding of some of the terminologies. The survey included questions on respondents’ demographic profile, interest in/history with FP, and their perceptions of and willingness to use and pay for a CFTA.

Participants were introduced to the concept of a CFTA via a 13-minute informational video. This video was designed to explain the method profile of a hypothetical app called “The Family Planning App.” While not an actual brand of CFTA, “The Family Planning App” depicted in the video comprised features of real apps on the market, including the effectiveness of those backed by clinical efficacy data (92% with typical use). The depicted app relies on the Standard Days Method of estimating effectiveness based on dates of menstruation, rather than biomarkers. This decision was made because apps requiring biomarkers often require additional tools, like a thermometer, or a level of health literacy that may be less accessible or prevalent in lower resource settings. The video introduced “The Family Planning App” as a new product and described its correct use, features, benefits, and limitations. It also shared information about the app in context by comparing and contrasting features of the app with other contraceptive methods available in the Philippines. The video was intended to ensure all respondents shared the same baseline understanding of how a CFTA fits into the wider method mix so that they could more accurately report on their interest in and intention to use this new method. The video was pretested and refined during the first qualitative phase of this study, the results of which are not included here. Following pretesting, the video was revised for clarity and ease of comprehension in response to participant feedback.

A sub-set of the online survey participants were followed up with in a subsequent IDI phase. The qualitative tool was developed based on the review of the first 200 responses from the online survey to gain further insight into trends and responses seen in the online surveys. The qualitative study aimed to gain further understanding of some of the overall response trends and observed patterns in the online survey. Respondents had an option of choosing between an English or Tagalog interview based on their preferred language.

Data collection for the online survey was initiated and completed in January 2023, and the qualitative IDIs were initiated and completed in March 2023.

### Data Management and Analysis

Qualitative Data: Audio recordings of the interviews were transcribed and translated into English. A general coding framework that included key study questions was developed prior to analysis of the qualitative data. We used an iterative, thematic approach to data analysis with the process being flexible enough to accommodate emerging themes. Emergent themes/sub-themes were discussed among the team and added or refined after consensus from the data analysis team. After the coding framework was finalized, two members coded the transcripts. Coded data were grouped together by the respective themes/sub-themes for a common understanding.

Quantitative Data: The quantitative data was analyzed using STATA 15.0. Data was cleaned for completeness, response logic, outliers, and missing values. Where dual current method use was reported, the method with highest efficacy was taken as current method used. During the data cleaning process, 43 participant records were dropped from the final data set due to inconsistent and/or inaccurate responses regarding current contraceptive method used. The analysis thereafter was conducted using a total of 1,557 respondents.

Descriptive analyses were conducted on the variables of interest. The outcome variable “interest in using the app” was generated based on the response to the 5-point Likert scale question: “If this product were available to you now, how interested would you be in using it now?” The responses of “Probably Want to Use” and “Definitely Want to Use” were combined to create an “Interested” variable, while “Definitely Do Not Want to Use”, “Probably Do Not Want to Use” and “Unsure” were combined to create a “Not Interested/Unsure” variable.

Logistic regression analysis was conducted to understand associations between the outcome variable, demographic variables, and several behavioral variables: intention to use FP in the future, current use of modern methods, likeliness to switch methods in the next 12 months, and use of a similar app (e.g., period tracking apps) in the past. Models were adjusted for demographic variables: residence, age (as a continuous variable), marital status, wealth quintiles, education, occupation, and parity, to generate the final model.

Wealth quintiles were calculated using tools and analysis approaches used by EquityTool.org. The online survey collected data on seven household asset ownership questions recommended for the Philippines, and the ownership data was multiplied by factor weights created using 2017 Philippines Demographic and Heath Survey’s urban wealth index.

Willingness to pay estimates for the CFTA were explored amongst respondents who were either interested in or currently unsure about using CFTA in the future. The survey tool included pre-determined prices ranging from USD 1 to USD 10 (equivalent in Philippine pesos) per month. For each respondent, a random starting point between 4-7 USD was assigned. They were asked if they were willing to pay a suggested price every month to use the app. If the response was “yes”, the tool was programed to ask if they would pay a higher stated price, increased by 1 USD for each positive response up to USD 10. At that point the respondent was asked the maximum price they would be willing to pay. If the response to the initial question was “no”, the tool asked whether they would be willing to pay a lower suggested price, decreasing by 1 USD for each negative response until less than 1 USD was reached. This methodology is shown in figure 2.

**Figure 2.**
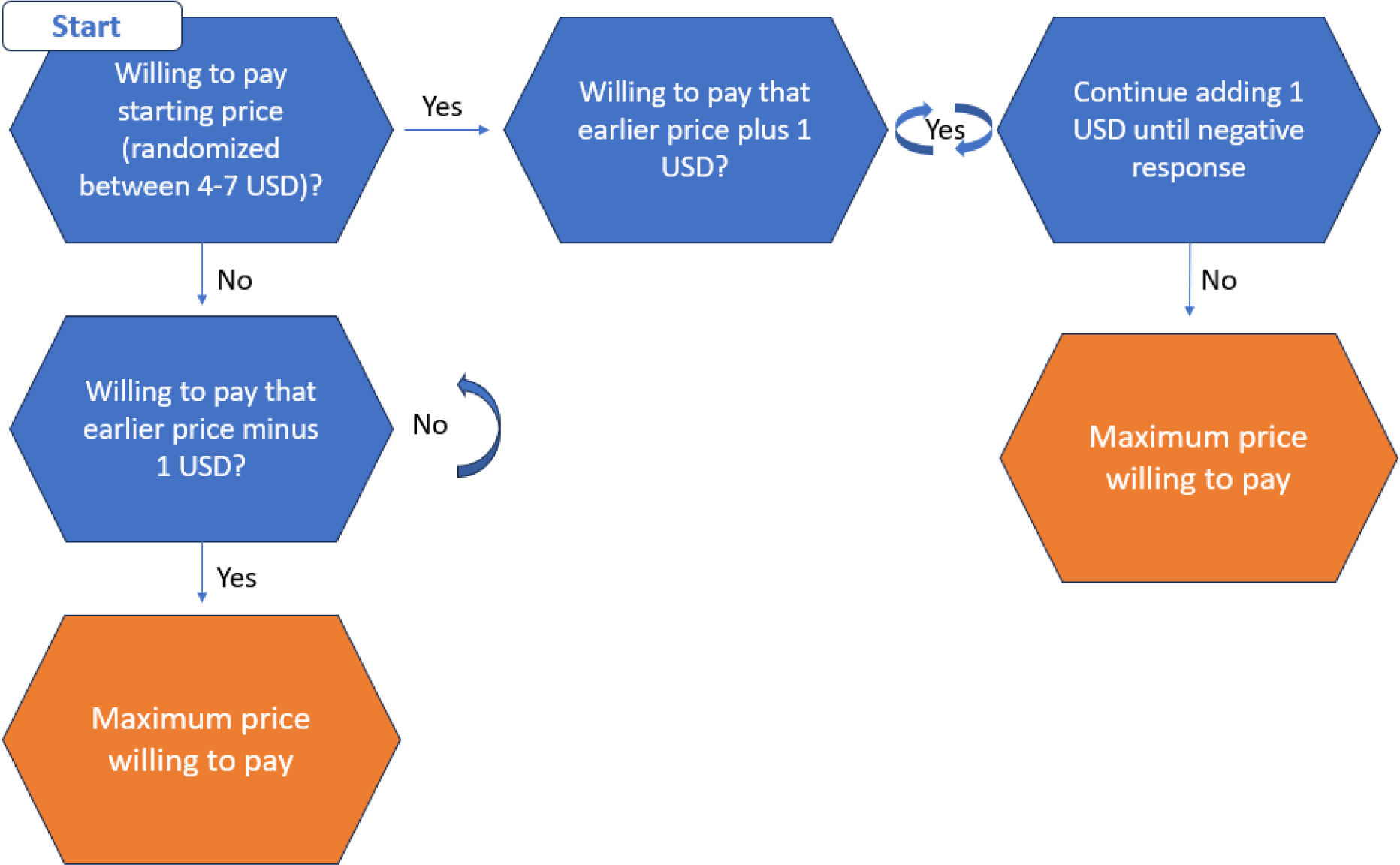
Willingness to Pay Methodology

### Ethics Approval

This study protocol and all associated data collection tools and forms were reviewed and approved by Population Services International’s (PSI) Research Ethics Board and the National Ethics Committee of the Philippines. Eligible respondents were provided with a description of the study and provided written (online) consent prior to participating in the study.

## Results

### Online Sample Characteristics

All participants for this survey were from urban areas, with the largest proportion (31%) from Metro Manila and the rest were distributed across urban areas of North & Central Luzon, South Luzon, Visayas, and Mindanao (Table 1). The sample was distributed similarly across different age groups in the study, with a mean age of 29.9 years. Over 70% of the participants were from the highest two wealth quintiles, a trend which may have been influenced by the nature of the panel and the selection criteria set for the study. Most participants were either married (46%) or in an exclusive relationship (40%). More than half (57%) of the participants had 2 or more children. Similarly, 47% had completed college and 52% were housewives.

**Table 1.** Percentage Distribution of Online Survey Respondents by Demographic Characteristics.

| **Demographic Characteristics** | **N=1557** |  |  |
| --- | --- | --- | --- |
|  | **Frequency (%)** |  | **Frequency (%)** |
| **Location** |  | **Age group** |  |
| Metro Manila | 487 (31.3) | 18-24yrs | 578 (37.1) |
| North & Central Luzon | 301 (19.3) | 25-34yrs | 484 (31.1) |
| South Luzon | 297 (19.1) | 35-49yrs | 495 (31.8) |
| Visayas | 237 (15.2) | *Median age (range)* | *29 yrs. (18-49)* |
| Mindanao | 235 (15.1) |  |  |
| **Wealth Quintiles (urban)** |  | **Relationship Status** |  |
| Lowest | 35 (2.6) | Married | 715 (45.9) |
| Second | 108 (6.9) | Exclusive Relationship, but Not Married | 615 (39.5) |
| Middle | 306 (19.7) | Single and Dating | 155 (10.0) |
| Fourth | 412 (26.5) | Single and Not Dating | 63 (4.1) |
| Highest | 696 (44.7) | I'd Rather Not Say | 9 (0.6) |
| **Number of Children** |  | **Education** |  |
| None | 299 (19.2) | Some Primary or Completed Primary (Completed Grade 6) | 33 (2.1) |
| 1 child | 378 (24.3) | Some Secondary | 153 (9.8) |
| 2+ children | 880 (56.5) | Completed Secondary<br>(Completed Grade 10) | 466 (29.9) |
|  |  | Post-Secondary | 173 (11.1) |
|  |  | College | 732 (47.0) |
| **Occupation** |  | **Current use for contraceptives** |  |
| Housewife | 813 (52.2) | Any Method | 1436 (92.2) |
| Professional | 209 (13.4) | Any Modern Method | 1181 (71.9) |
| Student/Working Student | 184 (11.8) | Oral Contraceptive Pills | 430 (27.6) |
| Currently Unemployed and Not a Student | 168 (10.8) | Male Condoms | 244 (15.7) |
| Other | 183 (11.8) | Injectable Contraceptive | 154 (9.9) |
|  |  | Implant | 84 (5.4) |
|  |  | Standard Days Method | 51 (3.3) |
|  |  | Emergency Contraceptive Pills | 45 (2.9) |
|  |  | Non-Hormonal IUD | 45 (2.9) |
|  |  | Hormonal IUD | 38 (2.4) |
|  |  | Other Modern Method | 29 (1.9) |
|  |  | Any Traditional Method | 252 (16.2) |
|  |  | Other Method (period tracking apps*) | 61 (3.9) |

\*Respondents reported using period tracking apps a contraceptive method (though they are not indicated for that purpose).

### Contraceptive Knowledge and Use

Nearly all participants (95%) had heard about at least one modern contraceptive method, with oral contraceptive pills (68%) and male condoms (68%) as the most frequently recalled modern methods. Similarly, 79% had ever heard about traditional methods of FP. One-third (37%) of the participants mentioned that they had ever heard about period tracking apps.

Current use of any FP method among participants was 92%, reflecting the selection criteria for the study. Among current users, most (71%) reported using a modern contraceptive method, with oral contraceptive pills (28%), male condoms (16%) and injectable contraceptive (10%) reported most often. Traditional method use was 16% and 4% were using a period tracking app (not a CFTA) exclusively as a contraceptive method.

Notably, dual method use was popular among the surveyed population with almost one in three (31%) reporting current use of more than one contraceptive method. A period tracking app was used by 8% of all current method users (dual and exclusive users) and nearly 23% of all participants had tried an app similar to “The Family Planning App” in the past. Among the period tracking app users (n=131), half mentioned using the app for spacing between children and 26% for delaying their first child. Online advertisements (48%) and referrals from a friend (33%) or family member (18%) were the most commonly mentioned sources of information on period tracking apps. Note that when asked to provide an app name, no respondents reported using one of the two apps that are currently approved for contraceptive use, therefore it is assumed that this does not indicate use of a CFTA.

Almost two-thirds of the participants mentioned they were “somewhat likely” or “very likely” to change their contraceptive method in the next 12 months. Only 17% said they are “unlikely” to change their current method in the next 12 months, while the rest were “undecided.”

### Interest in Using CFTA

Based on the information presented in the video, most participants (84%) reported they were “probably” or “definitely interested” in using a CFTA, if available (table 2). Comparatively, a higher proportion (19.2%) of the 18-24 age bracket were unable to decide on whether they would use a CFTA, if available. Similarly, 88% of participants reported they would recommend the app to others. Among those who “probably” or “definitely” wanted to use a CFTA, “helping to prevent pregnancy” (78%), “tracking menstrual cycle” (70%) and “tracking health changes or symptoms of their menstrual cycle” (63%) were the top reasons cited for interest.

**Table 2.** Reactions to the Video Information.

|  | **18-24**<br>**yrs.**<br>**N=578**<br>**(%)** | **25-34 yrs.**<br>**N=484 (%)** | **35-49**<br>**yrs.**<br>**N=495**<br>**(%)** | **Total**<br>**N=1557 (%)** |
| --- | --- | --- | --- | --- |
| **Interest in using app, if available (based on video)** |  |  |  |  |
| Definitely want to use it | 176<br>(30.5) | 245 (50.6) | 227<br>(45.9) | 648 (41.6) |
| Probably want to use it | 283<br>(49.0) | 172 (35.5) | 203<br>(41.0) | 658 (42.3) |
| Might or might not want to use it/not<br>able to decide now | 111<br>(19.2) | 63 (13.0) | 61 (12.3) | 235 (15.1) |
| Probably do not want to use it | 3 (0.5) | 1 (0.2) | 2 (0.4) | 6 (0.4) |
| Definitely do not want to use it | 5 (0.9) | 3 (0.6) | 2 (0.4) | 10 (0.6) |
| **Likelihood to recommend app to others (based on video)** |  |  |  |  |
| Strongly recommend this app | 193<br>(33.4) | 243 (50.2) | 225<br>(45.5) | 661 (42.5) |
| Recommend this app | 327<br>(56.6) | 181 (37.4) | 201<br>(40.6) | 709 (45.5) |
| Neutral/ I don't know | 45 (7.8) | 42 (8.7) | 53 (10.7) | 140 (9.0) |
| Don't recommend this app | 6 (1.0) | 2 (0.4) | 3 (0.6) | 11 (0.7) |
| Strongly don't recommend this app | 7 (1.2) | 16 (3.3) | 13 (2.6) | 36 (2.3) |
| **Reasons for wanting to use App<sup>a</sup> (%)** |  |  |  |  |
| Helping to prevent a pregnancy | 78.2 | 78.9 | 76.5 | 77.9 |
| Helping to track the days of your menstrual cycle | 75.8 | 66.9 | 65.6 | 69.6 |
| Helping to track health changes or symptoms of your menstrual cycle | 66.3 | 58.0 | 63.3 | 62.6 |
| Helping to plan a pregnancy | 51.9 | 46.8 | 42.1 | 47.0 |
| a. Multiple response. Among those who “probably” or “definitely wanted” to use a CFTA app (n=1306) |  |  |  |  |

Among those not showing interest in a CFTA (n=16), the most common reasons selected were “do not need contraception” (37%), “already have preferred method” (25%), and “irregular menstrual cycle” (25%). Similarly, among those who were “unsure” or “not able to decide” (n=235), 49% “needed to discuss with their partner,” 26% “wanted to discuss with a health provider,” and 25% “needed more time to decide if they wanted to use the app.”

### Correlates of App Use

We observed that the odds of residents of Visayas expressing interest in using the CFTA were lower compared to residents of Metro Manila (adjusted odds ratio [aOR]: 0.44; 95% confidence interval [CI]: 0.29-0.67) (table 3). For residents of other areas, including North & Central Luzon, South Luzon, and Mindanao, the odds of expressing interest were not significantly different as compared to Metro Manila residents. The odds of showing interest in using a CFTA also increased by 1.03 times with each unit increase in age (aOR: 1.03; 95% CI: 1.00-1.05). However, we did not observe significant differences in relation to other demographic variables.

**Table 3.** Interested in using App, if available.

|  | Unadjusted OR<sup>a</sup> | p-value | CI (95%) | Adjusted OR<sup>b</sup> | p-value | CI (95%) |
| --- | --- | --- | --- | --- | --- | --- |
| **Region (ref: Metro Manila)** |  |  |  |  |  |  |
| North & Central Luzon | 0.95 | 0.824 | 0.64-1.43 | 0.91 | 0.670 | 0.59-1.40 |
| South Luzon | 1.14 | 0.534 | 0.75-1.75 | 1.18 | 0.450 | 0.77-1.81 |
| Visayas | 0.45 | <0.001 | 0.31-0.67 | 0.44 | <0.001 | 0.29-0.67 |
| Mindanao | 0.99 | 0.973 | 0.64-1.55 | 1.09 | 0.711 | 0.68-1.77 |
| Age (continuous) | 1.02 | 0.025 | 1.00-1.04 | 1.03 | 0.039 | 1.00-1.05 |
| **Marital status (ref: currently married)** |  |  |  |  |  |  |
| Currently not married | 1.14 | 0.352 | 0.87-1.49 | 1.34 | 0.079 | 0.97-1.85 |
| **Wealth quintile (ref: lowest)** |  |  |  |  |  |  |
| Second | 0.98 | 0.972 | 0.40-2.44 | 0.92 | 0.854 | 0.36-2.33 |
| Middle | 1.35 | 0.482 | 0.58-3.14 | 1.17 | 0.731 | 0.49-2.80 |
| Fourth | 1.55 | 0.299 | 0.68-3.57 | 1.40 | 0.453 | 0.58-3.34 |
| Highest | 1.83 | 0.148 | 0.81-4.14 | 1.63 | 0.266 | 0.69-3.88 |
| **Education (ref: some secondary or lower)** |  |  |  |  |  |  |
| Completed secondary or post-secondary | 0.95 | 0.823 | 0.61-1.47 | 0.95 | 0.828 | 0.60-1.51 |
| College | 1.14 | 0.548 | 0.61-1.47 | 1.02 | 0.93<br>3 | 0.63-1.67 |
| **Occupation (ref: housewife)** |  |  |  |  |  |  |
| Student/working student | 1.16 | 0.531 | 0.73-1.82 | 1.60 | 0.09<br>6 | 0.92-2.79 |
| Currently unemployed | 0.70 | 0.088 | 0.46-1.06 | 0.74 | 0.16<br>2 | 0.48-1.13 |
| Professional | 1.54 | 0.073 | 0.96-2.47 | 1.68 | 0.06<br>2 | 0.97-2.89 |
| Other | 0.78 | 0.228 | 0.52-1.17 | 0.81 | 0.33<br>0 | 0.53-1.24 |
| **Parity (ref: none of 1 child)** |  |  |  |  |  |  |
| Two or more children | 1.12 | 0.415 | 0.85-1.47 | 1.15 | 0.40<br>5 | 0.83-1.60 |
| **Intend to use FP in future (ref: unsure)** |  |  |  |  |  |  |
| Yes | 2.68 | 0.009 | 1.28 -<br>5.59 | 2.55 | 0.01<br>9 | 1.17 -<br>5.58 |
| **Currently using modern method (ref: no)** |  |  |  |  |  |  |
| Yes | 1.36 | 0.038 | 1.02 -<br>1.81 | 1.39 | 0.03<br>5 | 1.02-1.90 |
| **Likely to begin new method in next 12 months (ref: no)** |  |  |  |  |  |  |
| Yes | 2.32 | <0.001 | 1.75 -<br>3.08 | 2.43 | <0.0<br>01 | 1.80 -<br>3.28 |
| **Used similar app in past (ref: no)** |  |  |  |  |  |  |
| Yes | 2.36 | <0.001 | 1.59 -<br>3.51 | 2.50 | <0.0<br>01 | 1.67 -<br>3.77 |
| a. OR = odds ratio<br>b. Adjusted for demographic variables: residence, age, marital status, wealth quintile, education, occupation, and parity. |  |  |  |  |  |  |

Odds of expressing interest in using the app were higher among those currently using a modern contraceptive method (aOR: 1.39; 95% CI: 1.02– 1.90) (table 3). Among current non-users, those who reported openness to use FP in the future had 2.6-fold odds of expressing interest in using a CFTA (aOR: 2.55;95% CI 1.17 - 5.58) relative to those not sure about using FP in the future holding age group, relationship status, wealth, education, parity, and occupation constant. Among current users, participants who reported that they were “probably” or “definitely” likely to begin using new contraceptive method in the next 12 months had higher odds of interest in the CFTA (aOR: 2.43; 95% CI: 1.80 - 3.28). Finally, participants who had used a similar (period tracking) app in the past had 2.5 times the odds of showing interest in using the CFTA, relative to those who had not used any similar app in the past (aOR: 2.50; 95% CI: 1.67 - 3.77).

### Willingness to Pay

We found that half (50%) of the survey participants had ever paid for their contraceptive method and 46% paid for their last contraceptive method used. The mean amount paid for their last contraceptive method was 237.33 Philippine pesos (USD 4.29) (n=722). The last purchased method for most participants was oral contraceptive pills (46%), followed by male condoms (24%), and contraceptive injectables (11%). While only 2% mentioned a period tracking apps as the method of contraception they had paid for last, 63% (n=1416) mentioned that someone like them would pay for a CFTA (data not shown). The price participants were willing to pay for the hypothetical app ranged from less than 58 pesos (USD 1.04) to a high of 1500 pesos (USD 27.12) per month. The mode price they were willing to pay was 290 pesos (USD 5.20). At this price point, it is estimated that 53% of the respondents would be willing to buy the CFTA (Figure 3). From the mode price of 290 pesos, there is a sharp decline in the proportion of people willing to buy as the price increases. When asked about payment preferences (n=1240), 61% mentioned that they would prefer to pay a smaller amount per month, 33% said a one-time payment would be preferable and 7% preferred an annual payment (data not shown).

**Figure 3.**
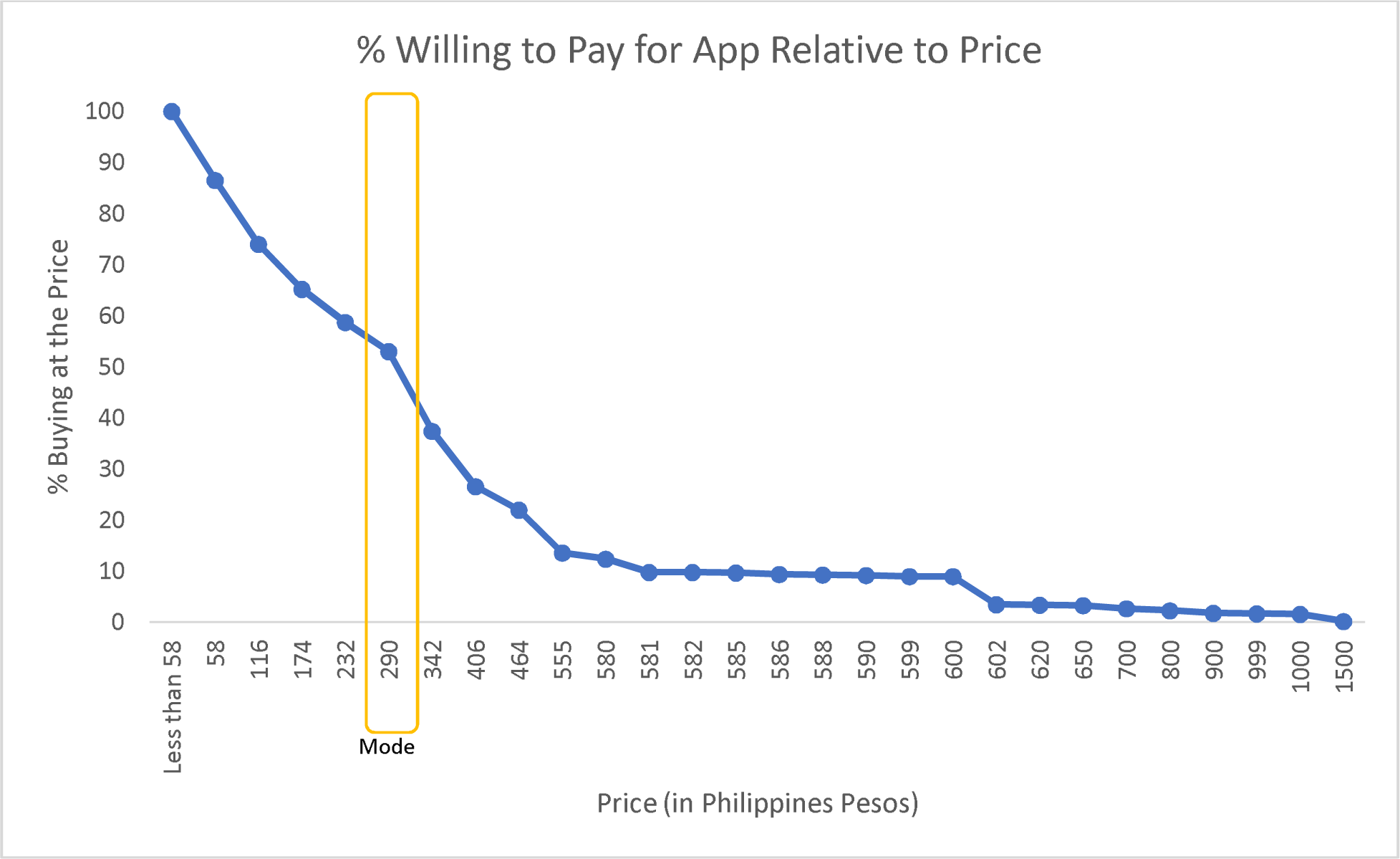
Willingness to Pay for CFTA

### Qualitative In-depth Interviews

When probed on the most appealing aspects of a hypothetical CFTA, the most common responses were that CFTAs were “natural” or “safe” and did not cause any side effects. Respondents also appreciated that CFTAs did not require visiting a health center to obtain, were easy to use and could even be used offline, which was a specifically cited benefit. As one respondent noted: “we control how long we want to use it or stop using it.” Additionally, women described the convenience of CFTAs due to the ubiquitous nature of smartphones; as reported by one respondent, “we bring our phones with us always.” The effectiveness of CFTAs was cited as an important characteristic as was the ability to use the app for multiple purposes (e.g., for pregnancy prevention as well as planning).

Opinions varied on whether it would be better to pay for a CFTA on a monthly versus annual basis, and in general, there was openness to paying for additional app features (i.e., in a premium version). Specifically, women reported interest in having access to telehealth consults via the app as well as features related to tracking moods and symptoms, fitness and health content, and information on linkages to local health centers and resources. When comparing the hypothetical app to existing period trackers available in the Philippines, respondents seemed willing to consider purchasing a subscription to the hypothetical app at a higher price if the features were deemed more valuable than what was available otherwise.

## Discussion and Implications

For the rapidly growing global population with access to a smartphone, the ability to learn about, download, and use a contraceptive method on one’s device - from anywhere in the world - presents an extraordinary opportunity to expand contraceptive access and autonomy.

When considering the addition of a new contraceptive method to a health market, there are many variables to consider: Ministries of Health need to weigh the benefits and repercussions to an overall health system; donors need to evaluate whether they can support or subsidize the cost of this new method; and FP program implementers need figure out how to achieve impact while meeting the diverse needs of users. However, evidence to inform their discussions and decisions is lacking. With little practical evidence available exploring the introduction of CFTAs, this research may offer insights to inform their considerations.

These findings begin to explore the potential market for CFTAs in the Philippines and indicate a high interest among an internet-engaged, urban population. Interest in using a CFTA was high across age ranges and demographic characteristics. However, we found that, relative to participants “unsure” or “not interested” in using a hypothetical CFTA, a higher proportion of those interested in using the app were older (>24 years older). The odds of being interested also improved among current FP users and those looking to begin using a new contraceptive method in the next 12 months.

One distinguishing characteristic of CFTAs is that in addition to contraception, they also offer non-contraceptive benefits to users, like the ability to track health changes, moods, sexual activity, etc., linkages to online information on health, relationships, and contraception, and the ability to switch to a “pregnancy planning” mode immediately should the user’s reproductive goals change. Indeed, many respondents showed an interest in using an app like this for more than just contraception and reported a willingness to pay more for these types of premium features. While this presents some challenges in measuring the market impact of CFTAs solely as contraception, it also represents an opportunity to serve a broader audience with a multipurpose technology that can meet their changing needs across a lifecycle. Expanding effective options for contraceptive methods that meet clients’ diverse needs is an important strategy for improving contraceptive uptake and satisfaction.

While apps that are intended as contraception are likely to incur some cost for download and/or monthly use, respondents appear willing to pay for an app at a price comparable to what they might pay for other contraceptive methods. In our study, more than half of respondents reported a willingness to pay a price of 290 pesos per month (USD 5.20). This price would likely require some level of subsidy from donors or special access pricing from app manufacturers, to ensure greater accessibility, which should be considered in any potential plans for introduction.

Clients were notably drawn to the appeal of CFTAs as a “safe” option, and the fact that it would be free from side effects that they had experienced or heard of with other methods was particularly important. They also highlighted the appeal of the app as “convenient” and appreciated that it could be used offline and without the need to visit a health provider. Convenience and ease of access also speaks to the potential benefits of CFTAs to a wider health system as self-care options that require fewer (or in this case, no) visits to a health provider could serve a larger market, overcome some supply-side barriers to choice and autonomy, and help to relieve an overburdened health system. The self-care nature of CFTAs, which allows women to seamlessly move between the different modes (e.g., pregnancy prevention, pregnancy planning, menstrual tracking), and start/stop usage as relevant to their life circumstances, is another benefit unique to CFTAs that can serve users across their FP journey.

Another factor in the potential success of CFTAs in new markets is the existence and ubiquity of other period tracking apps. In the Philippines, we found knowledge and use of period tracking apps to be prevalent and reported use of these apps as contraception was also fairly common (though they are not intended to be used for this purpose). While other methods of contraception certainly carve out a piece of a FP market, where many brands may be competitive or even where counterfeit products may encroach on market share, the case of CFTAs is unique. Given the numerous versions of period trackers available on the market that are not approved for contraceptive use, many of which are free to use, understanding what type of changes to the enabling environment — e.g., education, instruction, referral networks – would be necessary to help women use these more basic apps effectively. As of now, there is no clinical trial data available to understand whether simple period trackers could be used in a way that’s more efficacious than the Standard Days Method or traditional methods (like rhythm and withdrawal), though it is an avenue worth further exploration. Our results also suggest that the use of other apps is correlated with interest in using a CFTA, highlighting a potential gateway in the market to bridge the gap between the use of period trackers and CFTAs. Understanding the user perspective on the comparative advantage of a contraceptive app versus a basic period tracker would be a critical step toward understanding the potential impact of CFTAs.

### Strengths and Limitations

The data represents a sample from an online panel used in this research. While the Nielsen IQ online panel, used for various online panel marketing research, may be representative of the online population in the Philippines, the same cannot be said of the sample thus generalizability may be limited. The authors accepted this limitation of the online survey format primarily because of the comparative advantage of exploring interest in using and willingness to pay with the population considered most likely to be reached with a future introduction of a CFTA. It is difficult to say, however, exactly how interest in using a CFTA may differ among a rural population with smartphone access. For example, one could imagine that regular internet access may be scarce in rural areas, leading to less uptake of a CFTA. At the same time, however, a rural population may find it more challenging to access other contraceptive methods at a facility thus resulting in greater interest in an app that could be downloaded with data and used offline. In addition, an online approach to data collection allowed the research team to reduce in person contact with respondents during the COVID-19 pandemic.

The hypothetical nature of the CFTA presented to participants may limit their ability to reliably judge their willingness to use the app. Additionally, the way that this app was presented to participants (via an informational video) may not be how a client would realistically learn about a similar method. Since the introduction of an app like this is not currently planned, the promotion plan is unknown. While we wanted to ensure comprehensive information about the app was provided, potential users may, in a real introduction of the app, hear about the method via a social media advertisement that is more promotional in nature or by word of mouth, for example. These varied methods of promotion would likely impact the perceived appeal of the app, and therefore the willingness to use and pay for it.

## Conclusion

Findings from this mixed methods research study suggest that interest in a CFTA in the Philippines is high and, were an app to be introduced, the features of this natural and convenient method would appeal to users across demographic profiles. Though potential users report interest in the app, several factors should be considered when introducing a method like this into a FP market, including the promotion plan, competition from other available apps, and the potential for subsidized pricing in LMIC markets. While the impact of a method like CFTA could be hindered by the need for a smartphone, cost, and limited efficacy data, the potential for a CFTA to serve users across their reproductive lives with a method that can be accessed by anyone with a smartphone, anywhere in the world, broadens the potential of this method beyond the limits of currently available methods.

## Data Availability

All data for this manuscript will be available publicly in a de-identified format from the USAID Data Development Library (DDL).

https://nam11.safelinks.protection.outlook.com/?url=https%3A%2F%2Fdata.usaid.gov%2Fd%2F2s8u-n2vw&data=05%7C01%7Ckdanna%40psi.org%7C80176a14e0a7445f92b908db91eb290e%7Ccd9cb8ece621472a979a549ab5ba2470%7C0%7C0%7C638264211446003329%7CUnknown%7CTWFpbGZsb3d8eyJWIjoiMC4wLjAwMDAiLCJQIjoiV2luMzIiLCJBTiI6Ik1haWwiLCJXVCI6Mn0%3D%7C3000%7C%7C%7C&sdata=jzSRPb2QCaS0MpXz4qa9PgHbwY2wBLXUnuYR5PPOR2A%3D&reserved=0

https://data.usaid.gov/d/2s8u-n2vw

## Acknowledgements

The authors would like to thank the team at Nielsen IQ in the Philippines for their partnership and management of data collection for this research.

## References

1 Tran, N.T., Tappis, H., Moon, P. et al. Sexual and reproductive health self-care in humanitarian and fragile settings: where should we start?. Confl Health 15, 22 (2021). https://doi.org/10.1186/s13031-021-00358-5

2 WHO guideline on self-care interventions for health and well-being, 2022 revision. Geneva: World Health Organization; 2022.

3 Ross J, Stover J. Use of modern contraception increases when more methods become available: analysis of evidence from 1982–2009. Glob Health Sci Pract. 2013;. 1(2):203-212. http://dx.doi.org/10.9745/GHSP-D-13-00010.

4 Self-Care Trailblazers Group. 2022. Messaging Platform. https://media.psi.org/wp-content/uploads/2022/01/30233534/SCTG-Messaging-Platform_Jan-2022.pdf?_ga=2.28851900.1276059993.1688334350-1795504199.1677692113

5 Worsfold L, Marriott L, Johnson S, Harper JC. Period tracker applications: What menstrual cycle information are they giving women? Women’s Health (Lond). 2021 Jan-Dec;17:17455065211049905. doi: 10.1177/17455065211049905. PMID: 34629005; PMCID: PMC8504278.

6. Grand View Research. 2021. Women’s Health App Market Size and Share Report, 2030. https://www.grandviewresearch.com/industry-analysis/womens-health-app-market

7 Johnson S, Marriott L, Zinaman M. Can apps and calendar methods predict ovulation with accuracy?, Current Medical Research and Opinion. 2018, 34:9, 1587–1594, DOI: 10.1080/03007995.2018.1475348

8. The Performance of Fertility Awareness-based Method Apps Marketed to Avoid Pregnancy Marguerite Duane, Alison Contreras, Elizabeth T. Jensen, Amina White. The Journal of the American Board of Family Medicine. Jul 2016, 29(4) 508-511; DOI: 10.3122/jabfm.2016.04.160022

9 Victoria Jennings, Liya T. Haile, Rebecca G. Simmons, Jeff Spieler & Dominick Shattuck (2019) Perfect-and typical-use effectiveness of the Dot fertility app over 13 cycles: results from a prospective contraceptive effectiveness trial, The European Journal of Contraception & Reproductive Health Care, 24:2, 148–153, DOI: 10.1080/13625187.2019.1581164

10 Berglund Scherwitzl E, Lundberg O, Kopp Kallner H, Gemzell Danielsson K, Trussell J, Scherwitzl R. Perfect-use and typical-use Pearl Index of a contraceptive mobile app. Contraception. 2017 Dec;96(6):420–425. doi: 10.1016/j.contraception.2017.08.014. Epub 2017 Sep 4. PMID: 28882680; PMCID: PMC5669828.

11. Food and Drug Administration. “FDA allows marketing of first direct-to-consumer app for contraceptive use to prevent pregnancy”. August 10, 2018. https://www.fda.gov/news-events/press-announcements/fda-allows-marketing-first-direct-consumer-app-contraceptive-use-prevent-pregnancy

12. Lomas N. March 1, 2021. “Clue Gets FDA Clearance to Launch a Digital Contraceptive”. TechCrunch. https://techcrunch.com/2021/03/01/clue-gets-fda-clearance-to-launch-a-digital-contraceptive/

13 Victoria Jennings, Liya T. Haile, Rebecca G. Simmons, Jeff Spieler & Dominick Shattuck (2019) Perfect-and typical-use effectiveness of the Dot fertility app over 13 cycles: results from a prospective contraceptive effectiveness trial, The European Journal of Contraception & Reproductive Health Care, 24:2, 148–153, DOI: 10.1080/13625187.2019.1581164

14 Berglund Scherwitzl E, Lundberg O, Kopp Kallner H, Gemzell Danielsson K, Trussell J, Scherwitzl R. Perfect-use and typical-use Pearl Index of a contraceptive mobile app. Contraception. 2017 Dec;96(6):420–425. doi: 10.1016/j.contraception.2017.08.014. Epub 2017 Sep 4. PMID: 28882680; PMCID: PMC5669828.

15. Frequently Asked Questions. Natural Cycles. https://www.naturalcycles.com/faqs

16 Peragallo Urrutia, Rachel MD, MS; Polis, Chelsea B. PhD; Jensen, Elizabeth T. PhD; Greene, Margaret E. PhD; Kennedy, Emily MA; Stanford, Joseph B. MD, MSPH. Effectiveness of Fertility Awareness–Based Methods for Pregnancy Prevention: A Systematic Review. Obstetrics & Gynecology. 132(3):p 591-604, September 2018. | DOI: 10.1097/AOG.0000000000002784

17 Hough A, Bryce M. Exaggerating contraceptive efficacy: the implications of the Advertising Standards Authority action against Natural Cycles BMJ Sexual & Reproductive Health 2019;45:71–72.

18 Bull J, Rowland S, Lundberg O, et al. Typical use effectiveness of Natural Cycles: postmarket surveillance study investigating the impact of previous contraceptive choice on the risk of unintended pregnancy. BMJ Open 2019;9:e026474. doi: 10.1136/bmjopen-2018-026474

19 Victoria Jennings, Liya T. Haile, Rebecca G. Simmons, Jeff Spieler & Dominick Shattuck (2019) Perfect-and typical-use effectiveness of the Dot fertility app over 13 cycles: results from a prospective contraceptive effectiveness trial, The European Journal of Contraception & Reproductive Health Care, 24:2, 148–153, DOI: 10.1080/13625187.2019.1581164

20 Berglund Scherwitzl E, Lundberg O, Kopp Kallner H, Gemzell Danielsson K, Trussell J, Scherwitzl R. Perfect-use and typical-use Pearl Index of a contraceptive mobile app. Contraception. 2017 Dec;96(6):420–425. doi: 10.1016/j.contraception.2017.08.014. Epub 2017 Sep 4. PMID: 28882680; PMCID: PMC5669828.

21. Torchinsky R. June 24, 2022. “Reproductive Rights in America. How period tracking apps and data privacy fit into a post-Roe v. Wade climate” NPR https://www.npr.org/2022/05/10/1097482967/roe-v-wade-supreme-court-abortion-period-apps

22. Biswas R. March 10, 2023. Philippines amongst world’s fastest growing emerging markets. S&P Global: Market Intelligence. https://www.spglobal.com/marketintelligence/en/mi/research-analysis/philippines-amongst-worlds-fastest-growing-emerging-markets-mar23.html#:∼:text=The%20Philippines%20economy%20grew%20at,well%20as%20gross%20capital%20for mation

23. Pew Research Center, March 2019, “Mobile Connectivity in Emerging Economies”

24. FP2020 (2022). Philippines 2022 Indicator Summary Sheet. https://www.track20.org/Philippines

25. FP2020 (2022). Philippines 2022 Indicator Summary Sheet. https://www.track20.org/Philippines

26. UNFPA Philippines. (2021). Family Planning. https://philippines.unfpa.org/en/node/15304; Accessed: Sept 2021

27 Bertrand JT, Sullivan M, Knowles EA, Zeeshan MF, Shelton JD. Contraceptive method skew and shifts in method mix in low-and middle-income countries. Int Perspect Sex Reprod Health. 2014 Sep;40(3):144–53. doi: 10.1363/4014414. PMID: 25271650.

28. Philippine Statistics Authority (PSA) and ICF. 2018. Philippines National Demographic and Health Survey 2017. Quezon City, Philippines, and Rockville, Maryland, USA: PSA and ICF

